# Silent Drift and Site-level Variance in Clinical AI: Real-world performance and subgroup analysis for post-market monitoring of an FDA-cleared artificial intelligence (AI) triage tool for high-risk use case of pulmonary embolism

**DOI:** 10.64898/2026.11.10.26367739

**Authors:** Ketan Dayma, Callum Newton, Michal Kawzowicz, Palak Patel, Ryan Curl, Kent Ogden

## Abstract

**Background:** FDA-cleared artificial intelligence (AI) triage tool for pulmonary embolism (PE) detection has been widely used; however, its performance has not been analyzed for subgroups, clinical sites, and temporal stability.

**Methods:** This retrospective, six-site health system study evaluated the Aidoc BriefCase PE triage model (K190072) across 14,736 CT pulmonary angiography (CTPA) examinations between July 2023 and March 2026. Reference-standard PE characteristic labels were extracted from radiology reports using a zero-shot LLM pipeline validated against a radiologist-annotated 600 reports (κ =0.98). Model performance was evaluated across demographics, embolus characteristics, and site. Temporal behavior was evaluated on operating point by logistic regression, with and without site adjustment, and by segmented regression.

**Results:** PE was present in 1,402/14,736 examinations (9.5%). Overall sensitivity was 64.1% (95% CI, 61.5–66.6%), specificity 98.8% (95% CI, 98.6–99.0%), compared with 93% sensitivity and 93.7% specificity in the FDA 510(k) summary. Sensitivity fell with peripherality (central 88.6%, lobar or segmental 63.1%, subsegmental 33.3%) and with chronicity (acute 65.5%, chronic 57.4%). Sensitivity differed significantly between the two highest-volume sites, 68.7% and 55.7% (P < .001), with CT scanner as the only variable different between the two sites. The false-positive rate fell 40% per year (P<.001) at a similar rate at two sites, while sensitivity fell 15% per year at site A (P<.036), from 72.5% to 65.0% between two halves of the study period. The manufacturer did not report model updates or threshold changes during the study period.

**Conclusions:** Deployed sensitivity was substantially below the cleared benchmark and varied 13 points by site. The gradual shift in high specificity at the cost of low sensitivity, either from data drift or threshold change, either not monitored or not documented by the manufacturer, highlights the importance of independent continuous post-market monitoring.

## Introduction

Pulmonary embolism (PE) requires rapid and accurate diagnosis (Goldhaber & Bounameaux, 2012), as delayed treatment increases mortality (Smith et al., 2010). Computed tomography pulmonary angiography (CTPA) is the primary imaging modality, with increasing utilization contributing to strain on the radiology workforce (Kohli et al., 2017; Recht et al., 2020).

Artificial intelligence (AI)-enabled radiology triage tools are increasingly deployed to prioritize potentially critical examinations, with FDA clearance relying largely on validation with curated, retrospective datasets (Benjamens et al., 2020; Kelly et al., 2019; Muehlematter et al., 2021; Park & Han, 2018; Zech et al., 2018).

Although developmental studies report sensitivities exceeding 85% to 90%, post- deployment evaluations have documented lower sensitivity, particularly for chronic and peripheral emboli, with sensitivities as low as 63.0% (Cellina et al., 2025; Goldberg-Stein et al., 2026; Langius-Wiffen et al., 2023; Rothenberg et al., 2023; Shah et al., 2026; Weikert et al., 2020). Vendor-deployed operation analytics metrics do not capture false-negative rates or longitudinal changes in diagnostic performance, and thus, clinical outcomes.

Current FDA pathways do not require post-market deployment validation or continuous performance monitoring, and although the Predetermined Change Control Plan (PCCP) pathway has begun to address ongoing performance validation, more than 90% of devices cleared through this pathway do not report such monitoring mechanisms (Dayma et al., 2026).

However, gaps in longitudinal performance and site-level heterogeneity remain insufficiently characterized. Prior real-world evaluations have reported pooled across facilities and have not previously tracked the temporal trends of AI diagnostic metrics (Brady et al., 2024; Goldberg-Stein et al., 2026; Mansuri et al., 2026; Venugopal et al., 2026). We therefore evaluated the diagnostic performance of an FDA-cleared PE triage model across six sites of an academic health system over 33 months, stratifying by site and tracking performance month by month, to better evaluate the feasibility of independent institutional monitoring.

## Methods

### Study design and cohort

This retrospective, IRB-approved study (IRB 2420324-1) evaluated the post-deployment performance of BriefCase (Aidoc Medical, Tel Aviv, Israel, K190072)(U.S. Food and Drug Administration, 2024), an FDA-cleared convolutional neural network that flags CTPA examinations for PE in adults. AI inference logs were independently linked to finalized radiology reports across six sites of an academic health system from July 2023 through March 2026. Examinations with non-diagnostic reports or unreconciled accession numbers were excluded, yielding 14,736 examinations from 31,827 reports (Figure 2). Two high-volume sites, a university hospital (Site A) and a community hospital (Site B), contributed 14,560 examinations (98.8%).

**Figure 1:**
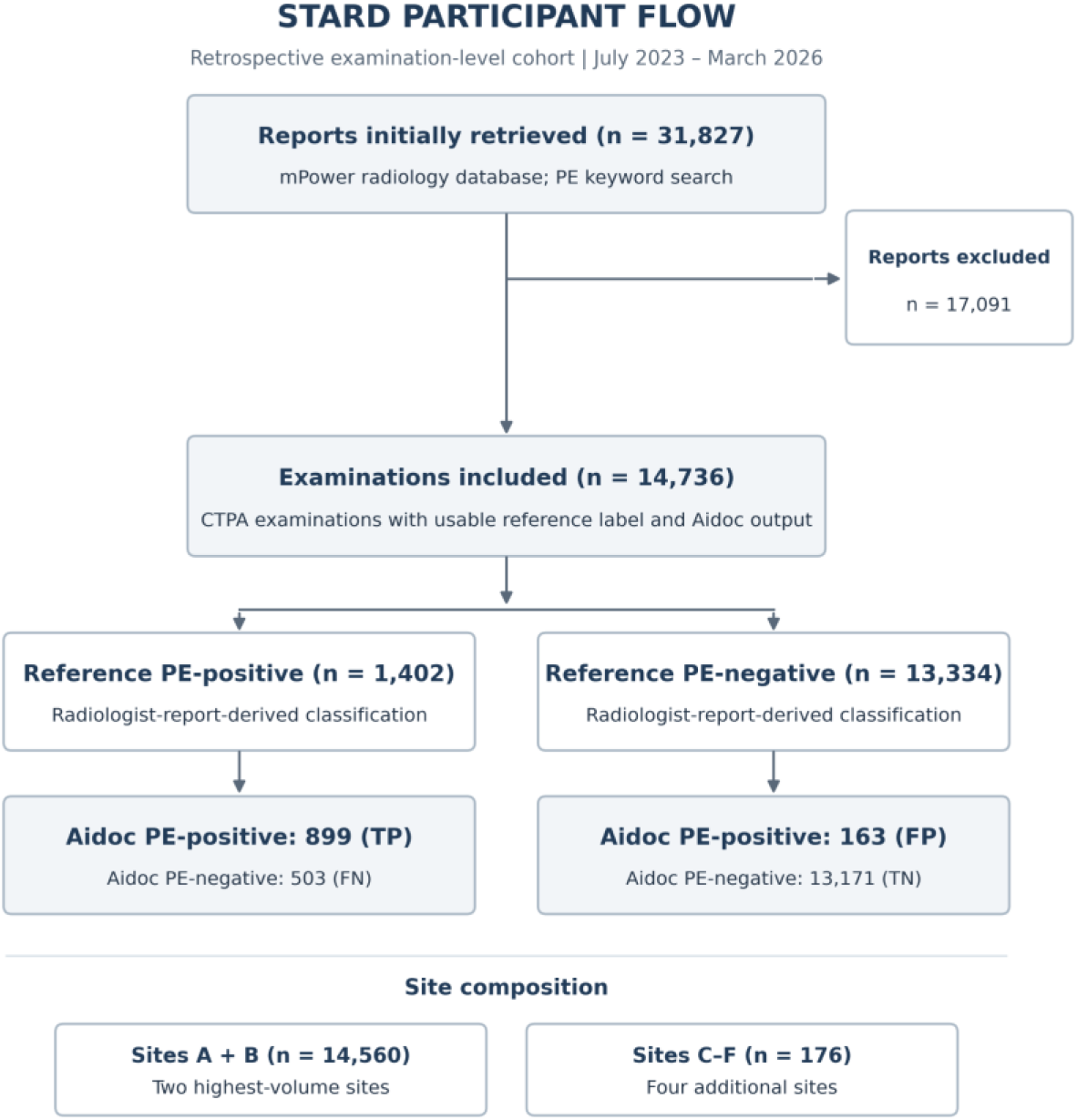
STARD case inclusion flow chart.

### Reference standard

A zero-shot prompting strategy with GPT-4o was used on a secure, HIPAA-compliant Azure deployment, mirroring methods validated for automated radiology text extraction in comparable large-scale studies (Goldberg-Stein et al., 2026; Mansuri et al., 2026; Sorin et al., 2025).

#### Primary detection

The LLM pipeline classified PE presence from the full text of 31,827 finalized reports spanning July 2023 to March 2026. The pipeline was instructed to parse negated phrasing and to prefer the impression over the findings section of the radiology report.

Because the AI device returns binary classifications without indeterminate state, equivocal reports were assigned reference-positive or reference-negative labels rather than excluded. Hedged negative statements which are often found in pre- specified templates at our institution (e.g., “small subsegmental embolus cannot be excluded”) were classified as negative, whereas hedged positive findings where radiologists explicitly added wordings (e.g., “could represent thrombus”) were classified as positive, according to our radiologist’s consensus (Table 1). Examinations reported as non-diagnostic were excluded.

**Table 1:**
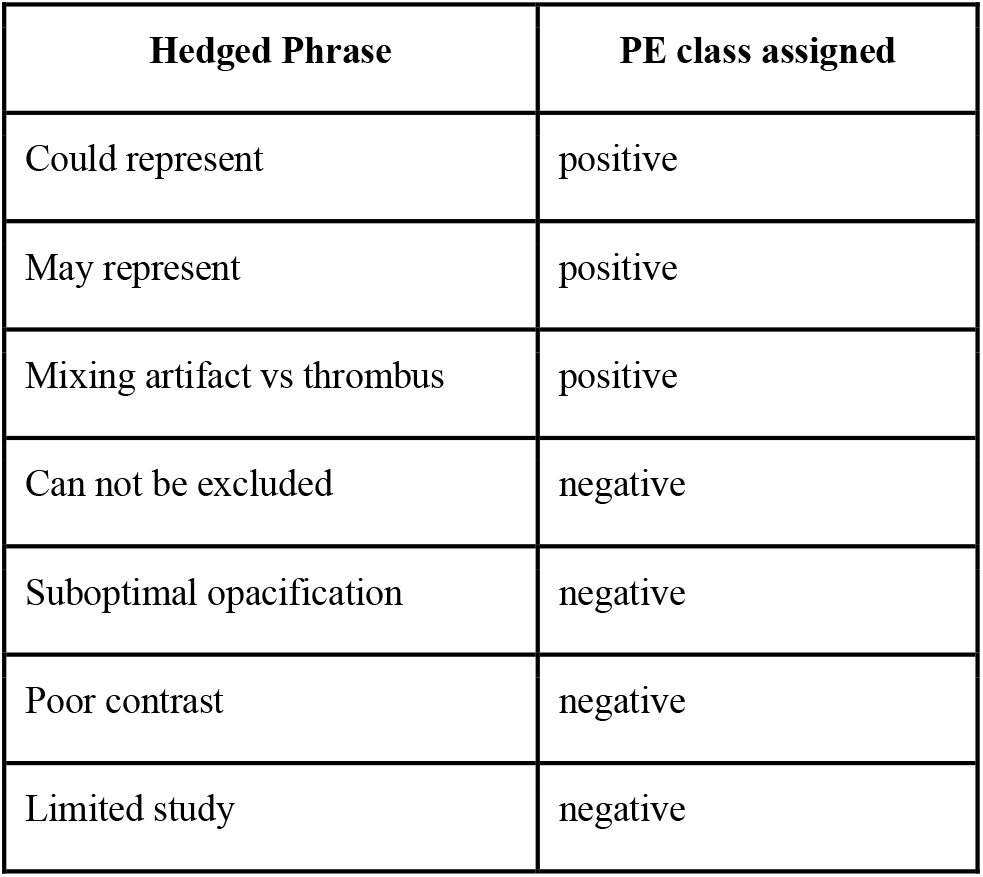
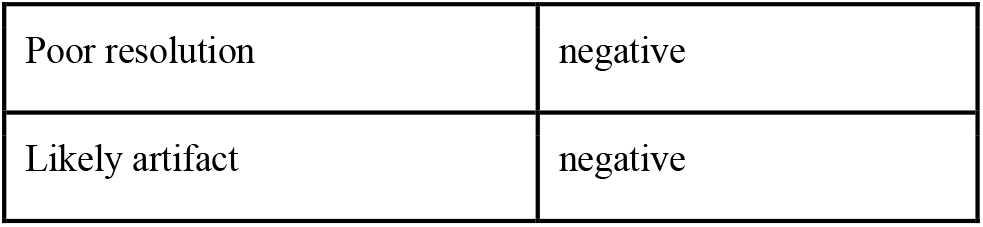
Hedged phrase lexicon used for negative or positive reference standards, based on radiologists’ consensus and institutional reporting practice.

| **Hedged Phrase** | **PE class assigned** |
|---|---|
| Could represent | positive |
| May represent | positive |
| Mixing artifact vs thrombus | positive |
| Can not be excluded | negative |
| Suboptimal opacification | negative |
| Poor contrast | negative |
| Limited study | negative |
| Poor resolution | negative |
| Likely artifact | negative |

Prompt performance was validated against an independent consensus reference standard established by a practicing radiologist and a PGY-5 radiology resident in 600 randomly selected reports (83 PE-positive, 517 PE-negative), reviewed independently and blinded to pipeline labels. Inter-reader agreement was 599/600 (Cohen’s κ = 0.99; 95% CI, 0.98–1.00). Extraction pipeline agreement was 597/600 (99.5%; 95% CI, 98.5–99.8%; κ = 0.98; 95% CI, 0.95–1.00), with 80/83 reference-positive (96.4%; 95% CI, 89.9–98.8%) and 517/517 reference-negative (100%; 95% CI, 99.3–100%) report agreement. All three cases of discordance were reference-positive examinations classified as negative by the pipeline.

#### Subgroup extraction

For the 1,402 examinations, additional prompts classified embolus chronicity and the most proximal arterial location. Validation was performed by a single annotator (a medical student trained by a radiologist) on 100 cases, demonstrating agreement rates of 96.0% for PE chronicity (Cohen’s κ = 0.696; 95% CI, 0.417–0.975) and 92.0% for PE location (κ = 0.885; 95% CI, 0.809–0.960). An exploratory analysis excluded 68 examinations with hedged- positive phrasing, retaining 1,334 definite-PE cases to assess sensitivity restricted to definite-positive examinations, consistent with prior comparator studies.

### Statistical analysis

Diagnostic performance was assessed using 2 × 2 contingency tables, with sensitivity, specificity, accuracy, predictive values, and false-positive and false-negative rates reported with 95% confidence intervals (Clopper-Pearson for pooled estimates; Wilson score for period-level estimates). Baseline characteristics were compared using Wilcoxon rank-sum and χ² tests, as appropriate.

Subgroup differences were evaluated using χ² or Fisher’s exact test with logistic regression and Holm-adjusted pairwise comparisons for clinical setting subgroups. Site-level differences were tested by logistic regression; sites contributing fewer than 100 examinations were reported descriptively and not hypothesis-tested.

Temporal change was evaluated separately with examination-level logistic regression modeling the odds of an AI-positive classification as a function of calendar month. Month-by-reference-status interaction tested differences between temporal slopes. Models were adjusted for site, with site-by-month interactions evaluating site-specific trends. Site-adjusted analyses were restricted to Sites A and B. Monthly heterogeneity and residual variation beyond linear trends were assessed using likelihood-ratio tests.

Segmented logistic regression evaluated potential change points, with significance assessed by parametric bootstrap. Early and late performance was compared using diagnostic odds ratios and Youden indices. CUSUM analyses assessed monitoring signals (Supplementary Methods). To evaluate changes in the case mix, quarterly distributions of embolus location, chronicity, technically limited examinations, and site volume were compared.

Statistical tests were two-sided (P < .05). Analyses were performed in R and Python using stats models. Reporting followed STARD 2015 guidelines; the checklist is provided in the Supplementary Material.

## Results

The cohort comprised 14,736 CTPA examinations, predominantly from emergency (70.7%) and inpatient (22.3%) settings. Median age was 63 years, 53.2% were female, and 84.0% of examinations were ordered STAT (Table 2). PE was identified in 1,402 examinations (9.5%). Most emboli were acute (88.2%) and involved lobar or segmental arteries (49.5%) (Table 3b).

**Table 2:** Demographic Subgroup Breakdown.

| **Characteristic** | **No. with data** | **Overall (N=14,736)** | **LLM FALSE (N=13,334)** | **LLM TRUE (N=1,402)** | **P value** |
|---|---|---|---|---|---|
| **Age, median (IQR), y, Median (Q1-Q3)** | 14,736 | 63.0 (49.0-73.0) | 63.0 (48.0-73.0) | 64.0 (50.0-74.0) | .004 |
| **Age group, No. (%), n (%)** | 14,736 |  |  |  | .11 |
| <18 |  | 1 (0.0%) | 1 (0.0%) | 0 (0.0%) |  |
| 18-65 |  | 8,360 (56.7%) | 7,609 (57.1%) | 751 (53.6%) |  |
| >65 |  | 6,375 (43.3%) | 5,724 (42.9%) | 651 (46.4%) |  |
| **Sex, No. (%), n (%)** | 14,735 |  |  |  | .01 |
| Female |  | 7,835 (53.2%) | 7,134 (53.5%) | 701 (50.0%) |  |
| Male |  | 6,900 (46.8%) | 6,199 (46.5%) | 701 (50.0%) |  |
| Missing |  | 1 | 1 | 0 |  |
| **Care setting, No. (%), n (%)** | 14,736 |  |  |  | <.001 |
| Emergency |  | 10,419 (70.7%) | 9,526 (71.4%) | 893 (63.7%) |  |
| Inpatient |  | 3,280 (22.3%) | 2,837 (21.3%) | 443 (31.6%) |  |
| Outpatient |  | 1,037 (7.0%) | 971 (7.3%) | 66 (4.7%) |  |
| **Order priority, No. (%), n (%)** | 14,736 |  |  |  | .45 |
| Routine |  | 2,362 (16.0%) | 2,127 (16.0%) | 235 (16.8%) |  |
| STAT |  | 12,374 (84.0%) | 11,207 (84.0%) | 1,167 (83.2%) |  |
| **Aidoc classification, No. (%), n (%)** | 14,736 |  |  |  | <.001 |
| FALSE |  | 13,674 (92.8%) | 13,171 (98.8%) | 503 (35.9%) |  |
| TRUE |  | 1,062 (7.2%) | 163 (1.2%) | 899 (64.1%) |  |

*Note.* Off-label processing of 1 erroneously acquired pediatric patient scan (<18 years) resulted in an uncalculable accuracy and a flat 50.0% artifact across remaining metrics due to severe sample size limitations.

### Overall Diagnostic Performance

Overall, sensitivity was 64.1% (899/1,402; 95% CI, 61.5–66.6%) and specificity was 98.8% (13,171/13,334; 95% CI, 98.6–99.0%). Accuracy was 95.5%, positive predictive value was 84.7%, and negative predictive value was 96.3% (Figure 2). Excluding 68 examinations with hedged-positive phrasing yielded a definite-PE sensitivity of 65.7% (95% CI, 63.1–68.2%), compared with 64.1% in the full cohort (Supplementary Table S1). No quarterly sensitivity estimate had a 95% confidence interval extending to the device’s 80% clearance performance goal (Table S4a, S4b).

**Figure 2:**
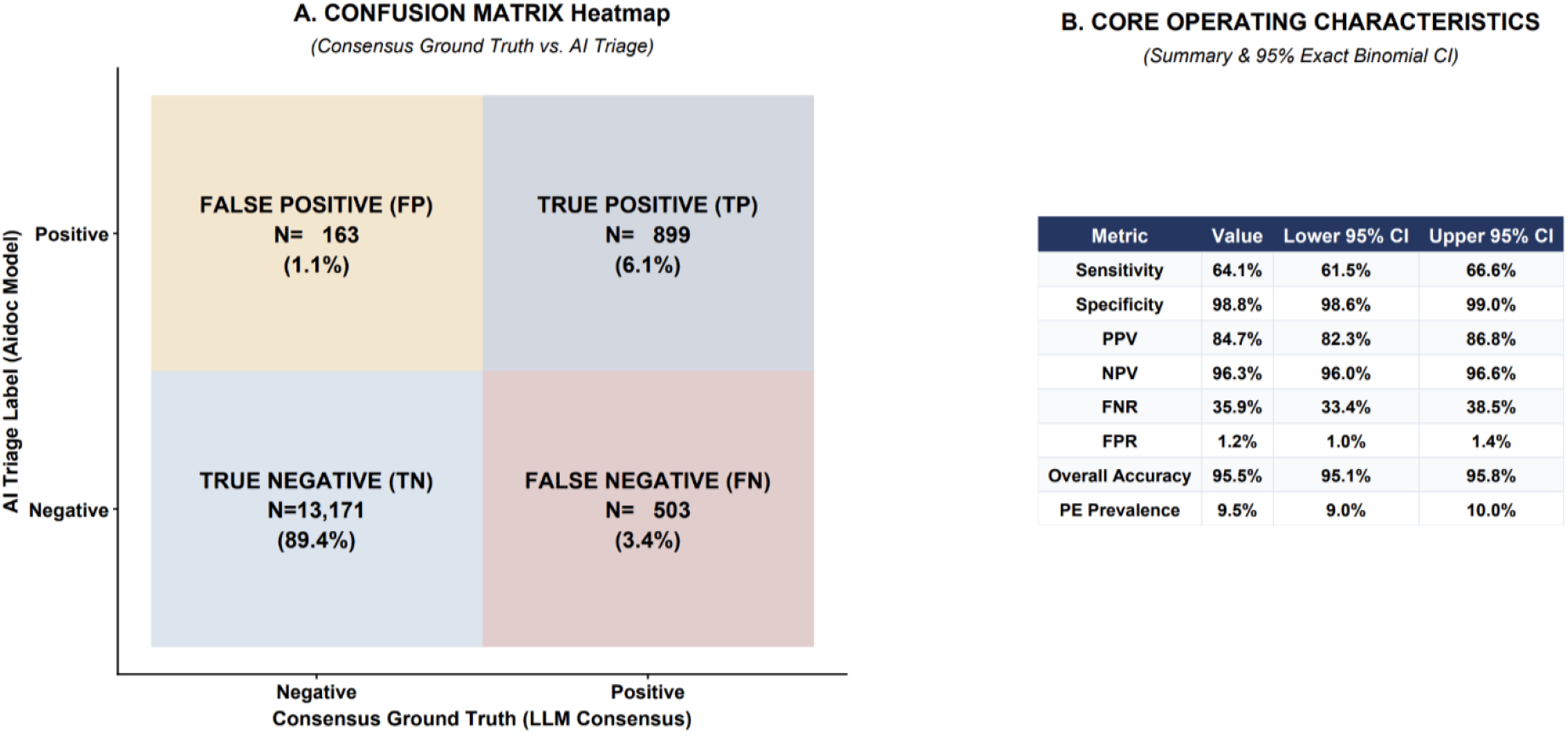
Aidoc Briefcase PE Model: Overall Headline Diagnostic Performance. *Note.* All statistics calculated using exact Clopper-Pearson binomial methods (Total Cohort N=14,736).

### Demographic, setting and STAT priority

No significant differences in sensitivity were observed by age or sex (Table 3a). Outpatient sensitivity was numerically lower (57.6%; 38/66) than inpatient (65.0%) or emergency sensitivity (64.2%), without significant heterogeneity (P = .50). Sensitivity did not differ significantly by order priority, although positive predictive value was higher for STAT than routine examinations (86.0% vs 78.9%; Table 3a).

**Table 3a:** AI performance by demographic stratification.

| Subgroup | Sensitivity | Specificity | Accuracy | PPV | NPV | FP rate | FN rate |
|---|---|---|---|---|---|---|---|
| **Overall** |  |  |  |  |  |  |  |
| Total population | 64.1% | 98.8% | 95.5% | 84.7% | 96.3% | 1.2% | 35.9% |
| **Patient sex** |  |  |  |  |  |  |  |
| Male | 65.9% | 98.7% | 95.4% | 85.2% | 96.2% | 1.3% | 34.1% |
| Female | 62.3% | 98.8% | 95.6% | 84.0% | 96.4% | 1.2% | 37.7% |
| **Age group** |  |  |  |  |  |  |  |
| 18–65 | 65.1% | 98.7% | 95.7% | 83.0% | 96.6% | 1.3% | 34.9% |
| >65 | 63.0% | 98.9% | 95.2% | 86.7% | 95.9% | 1.1% | 37.0% |
| **Clinical setting** |  |  |  |  |  |  |  |
| Emergency | 64.2% | 99.0% | 96.0% | 86.2% | 96.7% | 1.0% | 35.8% |
| Inpatient | 65.0% | 98.0% | 93.6% | 83.7% | 94.7% | 2.0% | 35.0% |
| Outpatient | 57.6% | 98.5% | 95.8% | 71.7% | 97.2% | 1.5% | 42.4% |
| **STAT status** |  |  |  |  |  |  |  |
| STAT | 63.6% | 98.9% | 95.6% | 86.0% | 96.3% | 1.1% | 36.4% |
| Non-STAT | 66.8% | 98.0% | 94.9% | 78.9% | 96.4% | 2.0% | 33.2% |

### Site-level variation

Sensitivity was higher at Site A (68.7%; 618/900) than at Site B (55.7%; 273/490) by a 13.0-percentage-point difference (OR for Site B vs Site A, 0.57; 95% CI, 0.45–0.72; P < .001). The four remaining sites each contributed fewer than 100 examinations and are reported descriptively: Site C 66.7% (73 examinations), Site D 60.0% (68), Site E 50.0% (32), and Site F 100% (7).

Site A had more inpatient, and Non-STAT cases, compared to Site B, which contained more Emergency and STAT cases. The main drivers of low sensitivity in Site B could be attributed to (surprisingly), its low performance in emergency and STAT subgroups (Table S3). Both sites used a standardized CTPA acquisition protocol and shared interpreting radiologists and had a grossly equal case mix between both sites for PE location and Chronicity, except for the CT scanner as the only different variable (Figure S4).

#### Embolus characteristics

Sensitivity declined with increasing peripherality, from 88.6% for central to 63.1% for lobar or segmental and 33.3% for subsegmental PE. Sensitivity was 65.5% for acute and 57.4% for chronic PE (Table 3b).

**Table 3b:** AI performance by embolus characterization.

| PE characteristic | No. with data | LLM TRUE (N=1,402) | Sensitivity |
|---|---|---|---|
| **Aidoc classification, No. (%), n (%)** | 1,402 |  | 64.1% |
| FALSE |  | 503 (35.9%) | - |
| TRUE |  | 899 (64.1%) | - |
| **PE anatomical location, No. (%), n (%)** | 1,402 |  | **-** |
| Central |  | 385 (27.5%) | 88.6% |
| Lobar or Segmental |  | 694 (49.5%) | 63.1% |
| Subsegmental |  | 231 (16.5%) | 33.3% |
| Indeterminate |  | 92 (6.6%) | 46.7% |
| **PE chronicity, No. (%), n (%)** | 1,402 |  | **-** |
| Acute |  | 1,236 (88.2%) | 65.5% |
| Chronic |  | 148 (10.6%) | 57.4% |
| Age Indeterminate |  | 18 (1.3%) | 22.2% |

#### Temporal change in the operating point

##### False-positive rate

Among 13,334 reference-negative examinations, the odds of an AI-positive classification declined progressively over time (OR per month, 0.958; 95% CI, 0.942–0.975; P < .001). Quarterly false-positive rate fell from 2.0% to 0.5% between Q3 2023 and Q1 2026, with specificity rising correspondingly from 98.0% to 99.5%, and positive predictive value from 75.4% to 92.4% (Figure 3). Declines were observed at both high-volume sites, without evidence of differing slopes (site-by-month interaction, P = .88). Between the first 18 and last 15 months, the false-positive rate fell from 1.75% to 0.67% at Site A (P < .001) and from 1.72% to 0.64% at Site B (P < .001).

**Figure 3:**
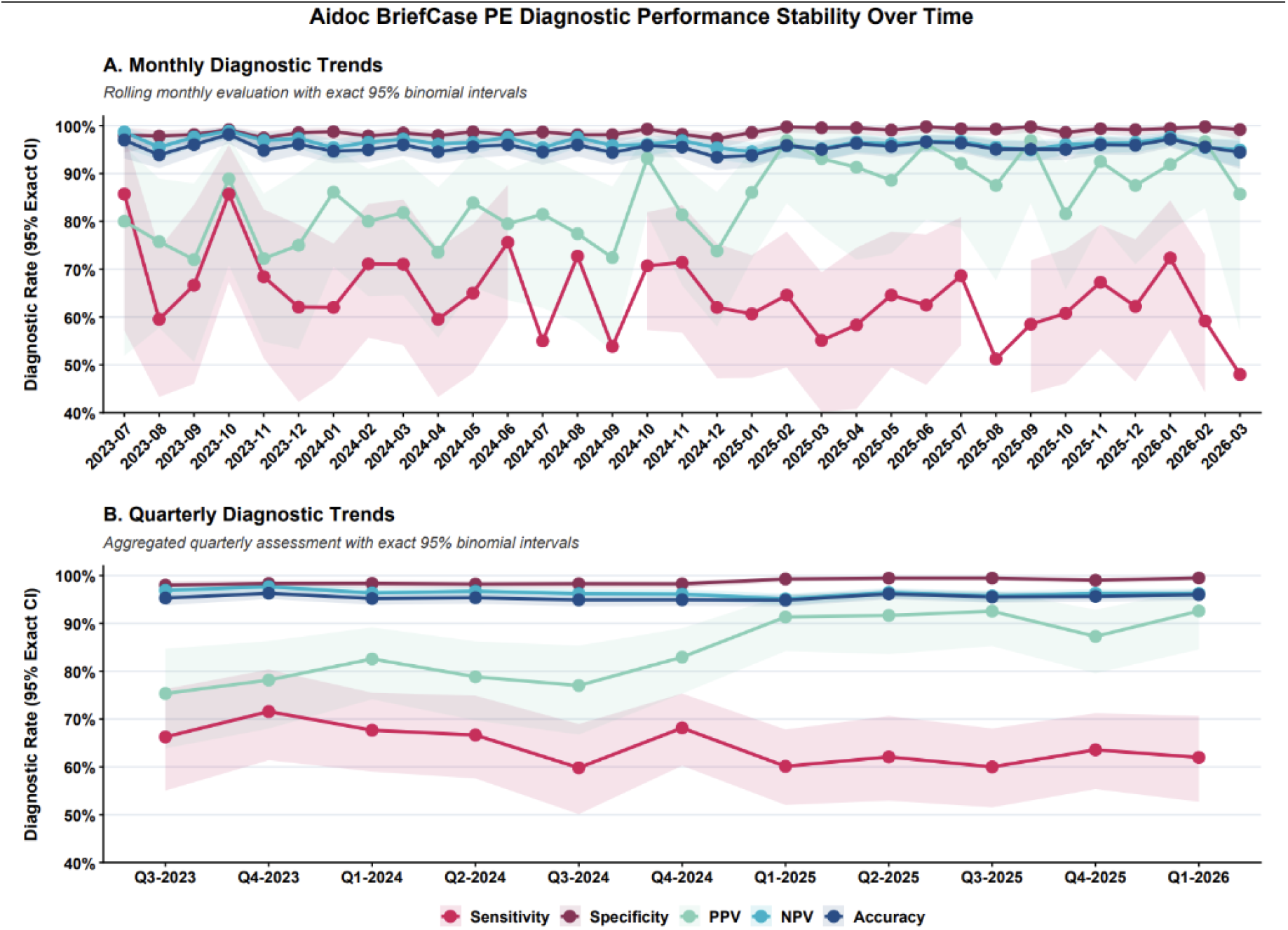
AIDoc Briefcase PE diagnostic performance stability over time.

##### Sensitivity

Among 1,402 reference-positive examinations, sensitivity declined non-significantly in the unadjusted analysis (OR per month, 0.988; 95% CI, 0.976– 1.000; P = .054), but significantly after site adjustment (OR, 0.987; 95% CI, 0.975–0.999; P = .036). At Site A, sensitivity decreased from 72.5% to 65.0% between the first 18 and last 15 months (P = .015), with a significant monthly trend (OR, 0.983; 95% CI, 0.968–0.998; P = .029). No significant trend was observed at Site B (P = .57), although the site-by-month interaction was nonsignificant (P = .38; Figure 3).

##### Difference between the two arms

The false-positive rate declined more steeply than sensitivity (month-by-reference-status interaction OR, 1.031; 95% CI, 1.010–1.053; P = .004), including after site adjustment (OR, 1.030; 95% CI, 1.009–1.053; P = .006). Between the early and late periods, the diagnostic odds ratio increased from 113 to 245, whereas the Youden index decreased from 0.650 to 0.609.

##### Change-point analysis

Segmented regression identified no significant system-level change point in either operating-point arm. A November 2024 change point in Site B’s false-positive rate reached nominal significance (bootstrap-adjusted P = .047; Supplementary Figures S2–S3).

##### Alternative explanations

Quarterly distributions of embolus location, chronicity, and hedged-positive reports showed no material changes. Monthly sensitivity estimates were imprecise at the study boundaries, including March 2026 (48.0%; 12/25; 95% CI, 30.0–66.5%). No software, algorithm, or threshold modification was communicated by the manufacturer.

## Discussion

In 14,736 CTPA examinations over 33 months, the AI triage model achieved 64.1% sensitivity (95% CI, 61.6–66.6%) and 98.8% specificity (98.6–99.0%), against the 93% sensitivity and 93.7% specificity reported in its 510(k) summary. Comparable drops in FDA clearance metrics for such models have now been described repeatedly: 86.8% across 30,678 CTPAs (Mansuri et al., 2026), approximately 85% across 32,501( Goldberg-Stein et al., 2026), and 82.2% for the same platform’s hemorrhage model across 101,944 head CTs against a cleared 96.2% (Chavoshi et al., 2025; Yu et al., 2022) found diminished performance in 81% of externally validated radiology algorithms. Therefore, the FDA clearance figures derived from relatively smaller validation studies should be considered as a ceiling, not an expectation.

Part of our lower estimate is methodological. Unlike the comparator studies, we retained examinations reported as limited, suboptimal, or possible. The device scores every study it receives and has no indeterminate state; technically limited examinations were scored anyway, and nearly always scored negative. A radiologist unaware of this behavior may assume the algorithm sees what a limited study obscures from the human eye. Restricting analysis to studies a radiologist judged diagnostic would measure something other than what clinicians encounter in real life. Excluding the 68 reference-positive cases resting on such hedged phrasing raised sensitivity only to 65.7%, with specificity unchanged.

Specificity exceeded the FDA clearance metric and stayed between 97.4% and 99.8% throughout, with a PPV of 84.7%. A flagged study was usually a true embolus, which limits alert fatigue and supports the tool’s intended role as a worklist triage. However, it does not support reading an unflagged study as evidence that PE is absent.

### Subgroup and Site Variation

The model worked best on acute proximal clot and degraded steadily as emboli became peripheral or older: 88.6% for central, 63.1% for lobar or segmental, 33.3% for subsegmental, and 57.4% for chronic versus 65.5% for acute (Mansuri et al., 2026). report the same gradient (99.3% saddle to 72.9% subsegmental), as does the hemorrhage literature for subacute and chronic bleeds. The likely explanation is as follows. Chronic emboli appear as isodense webs, flaps, or eccentric mural thrombus that a classifier may read as vessel wall or flow artifact, and peripheral vessels carry lower signal-to-noise with greater branching complexity (Nguyen et al., 2022; Wittram et al., 2004). Human readers show the same hierarchy; PIOPED II reported PPV falling from 97% for main or lobar emboli to 25% for subsegmental. Our false negatives concentrated in the territory where radiologist vigilance is already hardest to sustain and needs more assistance from such AI tools. Sensitivity was lower in outpatients (57.6%) than in the ED (64.2%) or inpatients (65.0%), although not statistically significant (p = 0.50, outpatient positives n = 66). Performance did not vary by age or sex.

The largest gap was between the facilities. Site A detected 68.7% of emboli and Site B 55.7%, a 13-point difference (OR 0.57; p < .001), despite sharing one acquisition protocol, 16-second contrast bolus timing, 120 kVp scans, a shared radiologist group, and equivalent distributions of embolus location and chronicity at both sites. From our physicist review, we identified one difference: Site A operates GE scanners, and Site B operates Siemens scanners. Image quality appears comparable to the eye, but reconstruction kernels and the compression applied before cloud transfer differ between vendors and either could plausibly alter the conspicuity of a small filling defect to a convolutional neural network (CNN). However, we cannot validate this from outside the device without collaboration with the manufacturer. Interestingly, the false-positive rate was identical at the two sites (1.23% and 1.20%), so whatever distinguishes Site B affects true emboli (high false negative rate) specifically rather than the handling of normal studies (low false positive rate). Neither of the two largest prior evaluations tested for between-site differences in sensitivity. Goldberg-Stein et al. compared hospitals with outpatient imaging centers, which is a comparison of facility types rather than two hospitals running matched protocols. This finding highlights the importance of site-level subgroup analysis, rather than one-time system-wide validation.

### Operative point shift versus Silent drift

Over 33 months, the false-positive rate fell by approximately 40% per year (p < .001) while sensitivity fell by approximately 15% per year in odds after site adjustment (p = .036), and the two slopes differed with statistical significance (p = .004). At Site A, this amounted to a fall from 72.5% to 65.0% between the first and second halves of the study. Such temporal change could be attributed to either local input data drift or AI-device-threshold change (Finlayson et al., 2021; Sahiner et al., 2023). The fall in sensitivity with rise in specificity is likely a sign of a moving decision to threshold. A true data drift should degrade both arms together.

Three additional observations argue against data drift. The false-positive decline was identical at both sites (OR 0.957 and 0.960; interaction p = .88), on different scanner platforms. Second, the study volume grew at the same rate at both sites between 2024 and 2025 (9.2% and 9.1%), so changing denominators cannot explain a sensitivity decline confined to Site A. Third, we did not find shifts over time in the distribution of embolus location or chronicity, and the same radiologists read throughout.

We cannot identify the true mechanism of such temporal change from outside the device. Incremental threshold adjustment, a phased model update, or a change in preprocessing upstream of inference would each produce this pattern. The manufacturer reported no version, algorithm, or threshold change during the study period. We could not verify that independently, despite our inquiry to the manufacturer, because version and threshold logs are not exposed to deploying sites and are not covered by our vendor agreement. It is important to note that the change was invisible through every channel available to us. The vendor analytics for this period report alert volumes and radiologist engagement of the tool, not the sensitivity against any independent reference standard, and so it could not have surfaced this, had this not been for our independent analysis.

### Clinical Implications

From the reading room, the rising specificity and fewer false alarms over time look like an improvement, inviting an automation bias. However, this came at the cost of low sensitivity: an unflagged study became steadily more likely to harbor an embolus. Nothing in the AI output or vendor communication signals this trade. A clinician who notices the high specificity will trust the tool more, not knowing the cost of the sensitivity arm. An emergency physician who sees an unflagged CTPA before the radiologist report may reasonably take it as reassurance; at a 36% false-negative rate, rising toward 50% in subsegmental and outpatient disease, the negative flag is not reassuring. This highlights the need for specialist-in-the-loop interpretation of unflagged examinations and the need for a defined “indeterminate” flag for non-evaluable studies or the certainty level of a binary flag to work as a safety control rather than a negative flag.

### Policy Implications

Our study highlights three important findings, worthy of shaping current policy on AI devices, being considered by the FDA (U.S. Food and Drug Administration, 2026). Monitoring triggered only by manufacturer-declared modification would not have triggered any monitoring here, because no modification was declared by the manufacturer; observed performance thresholds on a fixed cadence and subgroup analysis stratified by site are a necessary complement. A device cleared without an authorized PCCP sits in a regulatory gap: if a shift in our study reflects a vendor modification, the vendor alone judged whether it required a new submission for such modification, and if the temporal change reflects input data drift, such detection depends on post-market monitoring that FDA guidance recommends but does not mandate. Finally, the party best positioned to measure such a change; the vendor has the least incentive to disclose it. The deployed institution often lacks resources to do such monitoring at the level of expertise required (Chow et al., 2025; Sorin et al., 2025). Therefore, Independent evaluation by professional-society registries such as ACR Assess-AI, MDDT-qualified instruments, or third-party evaluator vendors is the only feasible arrangement in which measurement, incentive, and disclosure align, and its product should be a plain statement to end-users of how a device’s operating point has moved over time, to build trust from clinical end-users with AI devices.

### Limitations

First, this is a single health system study, with shared policies and a geographically homogeneous population, limiting generalizability. The reference standard was the finalized report, read with the AI result visible at the time of interpretation, so it cannot identify emboli missed by both the reader and the AI model. Labels were LLM-extracted; although agreement with two-reader consensus was 99.5% (κ = 0.98), residual label noise is likely, particularly in hedged phrasing or indeterminate location. Monthly denominators are small, and the first and last months were partial. The scanner hypothesis as a plausible cause cannot be tested from outside the black-box AI device. AI outputs from the vendor were made available up to March 2026, so we cannot say whether the trend continued thereafter.

### Conclusion

Deployed sensitivity was far below the FDA-cleared benchmark, varied by 13 points between two hospitals running identical protocols on different scanner platforms, and the sensitivity declined at the higher-performing site while the false-positive rate dropped at both. Whether this is vendor-level modification without communication of model update or true data drift between two sites, this study highlights the importance of independent post-market monitoring which cannot rest with the manufacturer or the deployed institution alone. Questions on who bears the liability when an FDA-cleared device changes behavior in use, as well as who carries the financial responsibility for such detection, remain open and are a priority for future work.

## Author contributions

KD and RC conceived and designed the study. KD and CN performed the primary data analysis and interpretation and drafted the original manuscript. PP contributed to data extraction, curation, and pooling, and assisted with data analysis and validation. MK contributed to manuscript editing, formatting, annotation, and critical revision for clarity and structure. RC and KO provided overall supervision, contributed to study design and interpretation, and critically reviewed the manuscript. All authors reviewed and approved the final manuscript and agree to be accountable for their respective contributions. KD and CN contributed equally as first authors.

## Funding

This research received no specific grant from any funding agency in the public, commercial, or not-for-profit sectors.

## Ethics

This retrospective study was approved by the SUNY Upstate Medical University Institutional Review Board (IRB 2420324-1) with a waiver of informed consent.

## Data availability

De-identified aggregate data supporting the findings, including per-month and per-site contingency tables, are provided in the Supplementary Material. Report-level data cannot be shared because they contain protected health information.

## Competing interests

No author has any financial relationship or competing interests with the manufacturer of the device evaluated.

## Use of AI in manuscript preparation

GPT-4o was used as a labeling instrument for reference-standard extraction, as described in Methods. Claude Opus 5.0 (Anthropic Inc.) was used for paraphrasing this manuscript to meet the journal’s word requirements.

## Notes

### Competing Interest Statement

The authors have declared no competing interest.

### Author Declarations

IRB of SUNY Upstate Medical University gave ethical approval for this work.

