## Supplemental Materials for "Silent Drift and Site-level Variance in Clinical AI: Real-world performance and subgroup analysis for post-market monitoring of an FDA-cleared artificial intelligence (AI) triage tool for high-risk use case of pulmonary embolism"

### Supplementary Material

**Table S1:** Diagnostic Performance of Aidoc for Pulmonary Embolism Detection after Exclusion of Indeterminate Cases.

| Reference-standard classification | Aidoc negative | Aidoc positive | Total |
|---|---|---|---|
| PE absent | 13,171 | 163 | 13,334 |
| PE present | 458 | 876 | 1,334 |
| **Total** | **13,629** | **1,039** | **14,668** |

Note: After exclusion of 68 examinations classified as indeterminate for PE, 14,668 examinations remained in the analysis. Of these, 1,334 were PE-positive, and 13,334 were PE-negative according to the LLM-derived reference standard. Aidoc correctly identified 876 PE-positive examinations (true positives) and 13,171 PE-negative examinations (true negatives), with 163 false positives and 458 false negatives. Sensitivity was 65.67%, specificity was 98.78%, accuracy was 95.77%, positive predictive value was 84.31%, and negative predictive value was 96.64%. The false-positive rate was 1.22%, and the false-negative rate was 34.33%.

**Figure S1:** Aidoc BriefCase PE Model: Site A Diagnostic Performance

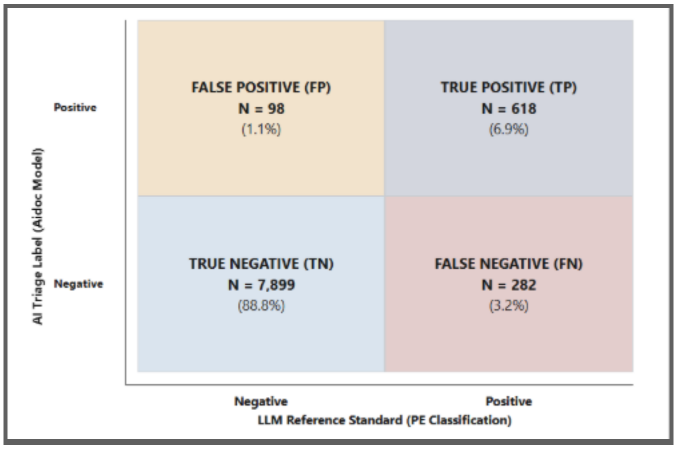


**Note.** Site A included 8,897 CTPA examinations, including 900 PE-positive cases (prevalence, 10.12%). Sensitivity was 68.67%; specificity, 98.77%; accuracy, 95.73%; positive predictive value (PPV), 86.31%; negative predictive value (NPV), 96.55%; false-positive rate (FPR), 1.23%; and false-negative rate (FNR), 31.33%. Percentages within heatmap cells indicate the proportion of the total campus cohort.

**Figure S2:** Aidoc BriefCase PE Model: Site B Diagnostic Performance

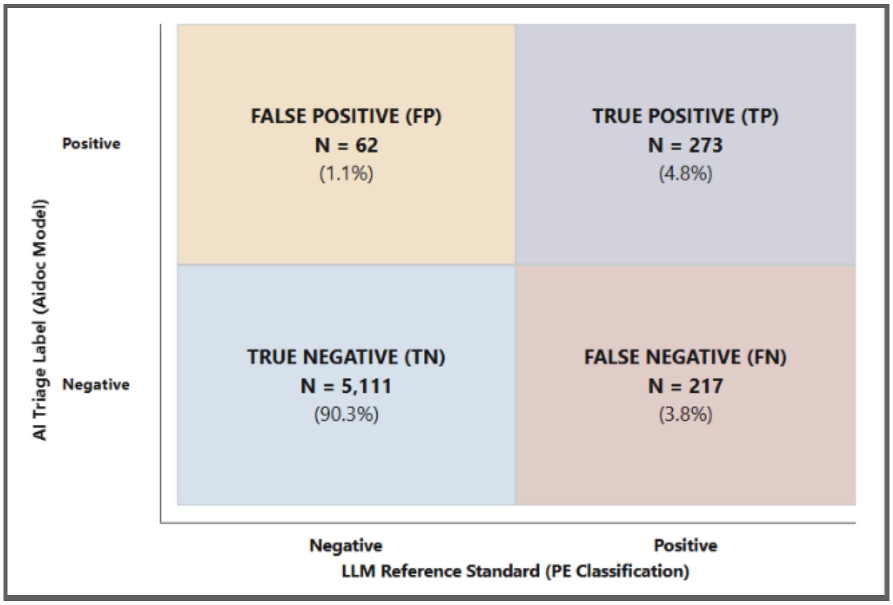


**Note.** Site B included 5,663 CTPA examinations, including 490 PE-positive cases (prevalence, 8.65%). Sensitivity was 55.71%; specificity, 98.80%; accuracy, 95.07%; positive predictive value (PPV), 81.49%; negative predictive value (NPV), 95.93%; false-positive rate (FPR), 1.20%; and false-negative rate (FNR), 44.29%. Percentages within heatmap cells indicate the proportion of the total campus cohort.

**Table S2a:** Site A diagnostic performance by clinical setting

| Clinical setting | N | Sensitivity (95% CI) | Specificity (95% CI) | Accuracy (95% CI) | PPV (95% CI) | NPV (95% CI) |
|---|---|---|---|---|---|---|
| All examinations | 8,897 | 68.7% (65.6–71.6) | 98.8% (98.5–99.0) | 95.7% (95.3–96.1) | 86.3% (83.6–88.6) | 96.6% (96.1–96.9) |
| Emergency | 5,847 | 70.9% (66.8–74.7) | 99.2% (98.9–99.4) | 96.8% (96.3–97.2) | 89.3% (85.9–92.0) | 97.4% (96.9–97.8) |
| Inpatient | 2,512 | 65.7% (60.7–70.3) | 97.7% (97.0–98.3) | 93.0% (91.9–93.9) | 83.3% (78.6–87.2) | 94.2% (93.2–95.1) |
| Outpatient | 538 | 68.8% (51.4–82.0) | 98.6% (97.2–99.3) | 96.8% (95.0–98.0) | 75.9% (57.9–87.8) | 98.0% (96.4–98.9) |

**Table S2b:** Site B diagnostic performance by clinical setting

| Clinical setting | N | Sensitivity (95% CI) | Specificity (95% CI) | Accuracy (95% CI) | PPV (95% CI) | NPV (95% CI) |
|---|---|---|---|---|---|---|
| All examinations | 5,663 | 55.7% (51.3–60.1) | 98.8% (98.5–99.1) | 95.1% (94.5–95.6) | 81.5% (77.0–85.3) | 95.9% (95.4–96.4) |
| Emergency | 4,572 | 55.8% (50.9–60.6) | 98.8% (98.4–99.1) | 95.1% (94.4–95.6) | 81.6% (76.6–85.8) | 95.9% (95.3–96.5) |
| Inpatient | 768 | 61.4% (49.7–72.0) | 99.0% (97.9–99.5) | 95.6% (93.9–96.8) | 86.0% (73.8–93.0) | 96.2% (94.6–97.4) |
| Outpatient | 323 | 36.4% (19.7–57.0) | 98.3% (96.2–99.3) | 94.1% (91.0–96.2) | 61.5% (35.5–82.3) | 95.5% (92.6–97.3) |

**Table S2c:** Site A vs. B Sensitivity by PE chronicity and anatomical location

| PE characteristic | Site A (N) | Site A sensitivity (95% CI) | Site B (N) | Site B sensitivity (95% CI) |
|---|---|---|---|---|
| **PE chronicity** | | | | |
| Acute | 775 | 70.3% (67.0–73.4) | 454 | 57.0% (52.5–61.5) |
| Chronic | 111 | 63.1% (53.8–71.5) | 32 | 40.6% (25.5–57.7) |
| Age indeterminate | 14 | 21.4% (7.6–47.6) | 4 | 25.0% (4.6–69.9) |
| **Anatomical PE location** | | | | |
| Central | 251 | 89.6% (85.3–92.8) | 130 | 86.2% (79.2–91.1) |
| Lobar/segmental | 449 | 67.9% (63.5–72.1) | 240 | 54.6% (48.3–60.8) |
| Subsegmental | 139 | 40.3% (32.5–48.6) | 91 | 22.0% (14.7–31.5) |
| Indeterminate | 61 | 52.5% (40.2–64.5) | 29 | 34.5% (19.9–52.7) |

**Campus Case-Mix Analysis**

Site A vs. B | PE-positive examinations | July 2023–March 2026

The master dataset provided all CTPA characteristics; the PE Characterization dataset supplied PE phenotype and Aidoc outcomes. No patient-level linkage was attempted. Analysis included 900 PE-positive Site A and 490 PE-positive Site B cases (n=1,390); 12 PE-positive cases from other sites were excluded from campus comparisons. Time was represented by calendar month, with no time-of-day analysis. PE-positive logistic models compared correct Aidoc detection at Site A versus Site B. Sequential models added calendar month, PE location/chronicity, and setting/STAT/age/sex. Model-based direct standardization used the pooled covariate distribution, with stratified campus bootstrap (400 attempts; 395 successful). Subgroup sensitivity used Wilson 95% confidence intervals and Fisher tests with exploratory Holm adjustment.

The Site A and Site B populations differ significantly in PE chronicity, clinical setting, STAT status, age, and sex, but not in overall PE anatomical location distribution. Site A sensitivity was 68.7% (618/900), compared with 55.7% (273/490) at Site B. The observed 13.0-percentage-point difference persisted after standardization to a common measured case mix (13.8 percentage points; bootstrap 95% CI, 8.8– 19.2). Thus, the measured covariates do not explain away the campus performance gap. This is an association and does not establish intrinsic model drift or a campus causal effect.

**Table S3:** PE-positive case-mix comparison

| Characteristic | Site A (N = 900) | Site B (N = 490) | P value |
|---|---|---|---|
| **PE location** | | | 0.4747 |
| Central | 251 (27.9%) | 130 (26.5%) | |
| Lobar or segmental | 449 (49.9%) | 240 (49.0%) | |
| Subsegmental | 139 (15.4%) | 91 (18.6%) | |
| Indeterminate | 61 (6.8%) | 29 (5.9%) | |
| **PE chronicity** | | | 0.0013 |
| Acute | 775 (86.1%) | 454 (92.7%) | |
| Chronic | 111 (12.3%) | 32 (6.5%) | |
| Age-indeterminate | 14 (1.6%) | 4 (0.8%) | |
| **Clinical setting** | | | **<0.0001** |
| Emergency department | 495 (55.0%) | 398 (81.2%) | |
| Inpatient | 373 (41.4%) | 70 (14.3%) | |
| Outpatient | 32 (3.6%) | 22 (4.5%) | |
| **STAT status** | | | **<0.0001** |
| Positive | 181 (20.1%) | 47 (9.6%) | |
| Negative | 719 (79.9%) | 443 (90.4%) | |
| **Sex** | | | 0.0003 |
| Female | 415 (46.1%) | 276 (56.3%) | |
| Male | 485 (53.9%) | 214 (43.7%) | |

Note. PE location distribution did not differ significantly (p=0.475). Chronicity did (p=0.0013): Site A had more chronic PE (12.3% versus 6.5%), whereas Site B had more acute PE (92.7% versus 86.1%). ED cases represented 81.2% of Site B PE-positive examinations versus 55.0% at Site A; inpatient cases were 14.3% versus 41.4%. These are distributional tests; repeated p-values across category rows reflect the same overall test, not separate category-level hypothesis tests.

**Figure S3:** Campus sensitivity by PE phenotype and clinical subgroup

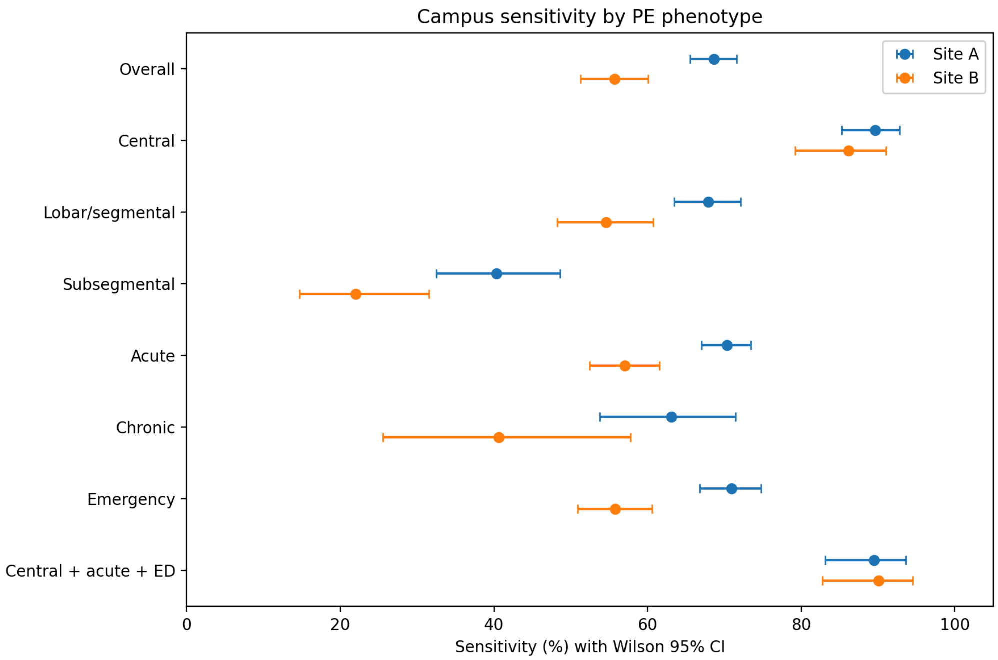


Note. Points show observed sensitivity for Sites A and B, and horizontal bars indicate Wilson 95% confidence intervals. The figure includes overall PE-positive cases and selected strata by anatomical location, chronicity, emergency setting, and the combined central/acute/ED subgroup. Site A sensitivity was higher for lobar/segmental and subsegmental PE, whereas central/acute/ED sensitivity was similar between campuses.

**Figure S4:** Distribution of PE location and chronicity among PE-positive examinations at Sites A and B

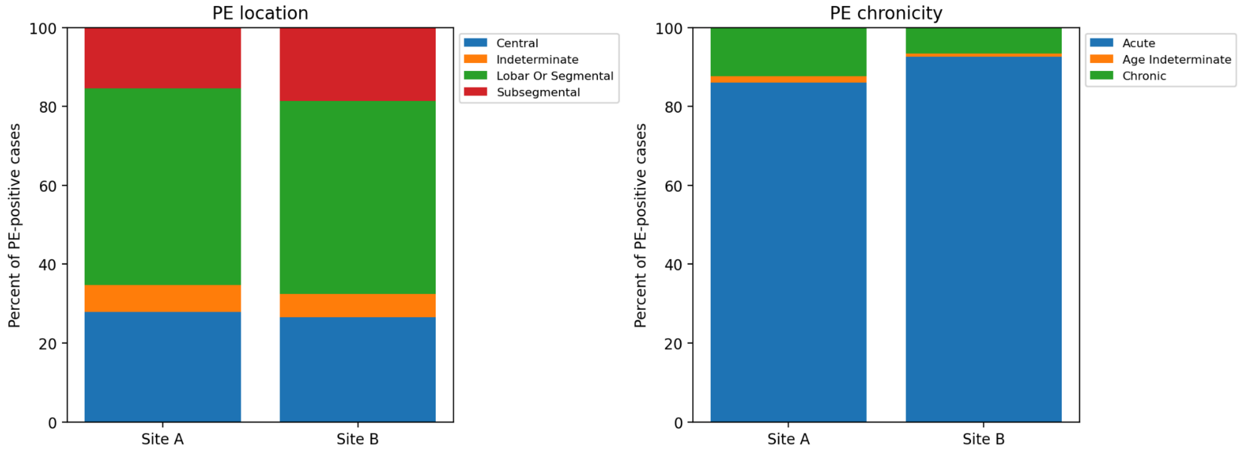


Note. Stacked bars show the percentage distribution of PE anatomical location (left) and chronicity (right) within each campus. The overall location distribution did not differ significantly (chi-square p=0.475), despite a numerically greater proportion of subsegmental PE at CC. Chronicity distribution differed (p=0.0013), with a greater chronic PE share at Site A (12.3% versus 6.5%) and a greater acute PE share at CC (92.7% versus 86.1%).

**Figure S5:** Longitudinal Monitoring of Sensitive Operating Characteristics

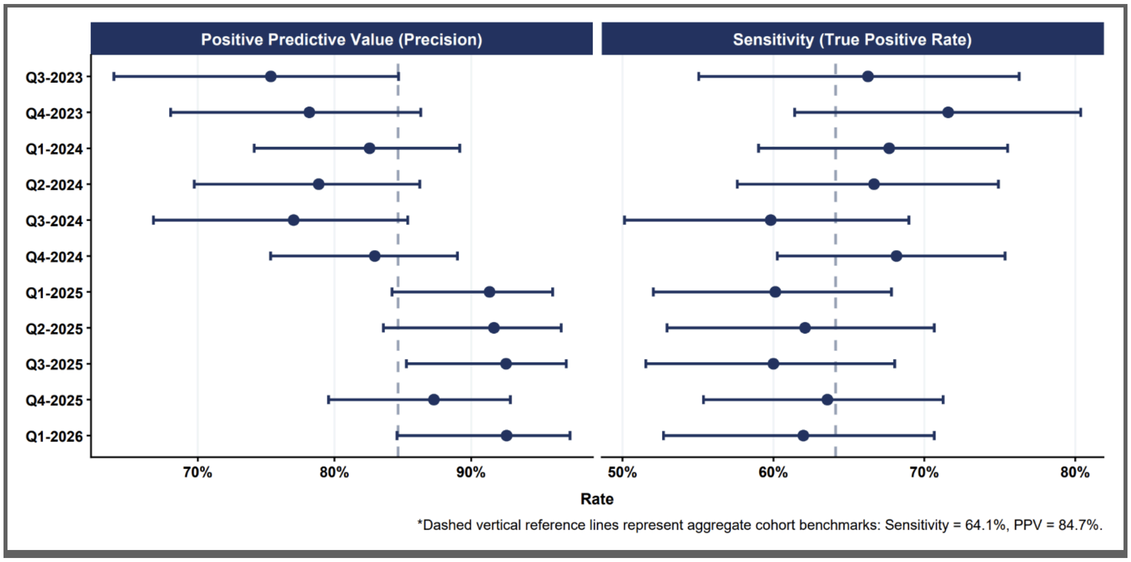

#### Table S4a**:** Monthly temporal stability and performance drift

| Period | Total N | PE positive, No. (%) | PE negative, No. (%) | Sensitivity (95% CI) | Specificity (95% CI) | Accuracy (95% CI) |
|---|---|---|---|---|---|---|
| Jul 2023 | 167 | 14 (8.4%) | 153 (91.6%) | 85.7% (60.1%–96.0%) | 98.0% (94.4%–99.3%) | 97.0% (93.2%–98.7%) |
| Aug 2023 | 409 | 42 (10.3%) | 367 (89.7%) | 59.5% (44.5%–73.0%) | 97.8% (95.8%–98.9%) | 93.9% (91.1%–95.8%) |
| Sep 2023 | 404 | 27 (6.7%) | 377 (93.3%) | 66.7% (47.8%–81.4%) | 98.1% (96.2%–99.1%) | 96.0% (93.7%–97.5%) |
| Oct 2023 | 382 | 28 (7.3%) | 354 (92.7%) | 85.7% (68.5%–94.3%) | 99.2% (97.5%–99.7%) | 98.2% (96.3%–99.1%) |
| Nov 2023 | 426 | 38 (8.9%) | 388 (91.1%) | 68.4% (52.5%–80.9%) | 97.4% (95.3%–98.6%) | 94.8% (92.3%–96.6%) |
| Dec 2023 | 436 | 29 (6.7%) | 407 (93.3%) | 62.1% (44.0%–77.3%) | 98.5% (96.8%–99.3%) | 96.1% (93.8%–97.6%) |
| Jan 2024 | 449 | 50 (11.1%) | 399 (88.9%) | 62.0% (48.2%–74.1%) | 98.7% (97.1%–99.5%) | 94.7% (92.2%–96.4%) |
| Feb 2024 | 416 | 45 (10.8%) | 371 (89.2%) | 71.1% (56.6%–82.3%) | 97.8% (95.8%–98.9%) | 95.0% (92.4%–96.7%) |
| Mar 2024 | 427 | 38 (8.9%) | 389 (91.1%) | 71.1% (55.2%–83.0%) | 98.5% (96.7%–99.3%) | 96.0% (93.7%–97.5%) |
| Apr 2024 | 476 | 42 (8.8%) | 434 (91.2%) | 59.5% (44.5%–73.0%) | 97.9% (96.1%–98.9%) | 94.5% (92.1%–96.2%) |
| May 2024 | 431 | 40 (9.3%) | 391 (90.7%) | 65.0% (49.5%–77.9%) | 98.7% (97.0%–99.5%) | 95.6% (93.2%–97.2%) |
| Jun 2024 | 451 | 41 (9.1%) | 410 (90.9%) | 75.6% (60.7%–86.2%) | 98.0% (96.2%–99.0%) | 96.0% (93.8%–97.5%) |
| Jul 2024 | 418 | 40 (9.6%) | 378 (90.4%) | 55.0% (39.8%–69.3%) | 98.7% (96.9%–99.4%) | 94.5% (91.9%–96.3%) |
| Aug 2024 | 398 | 33 (8.3%) | 365 (91.7%) | 72.7% (55.8%–84.9%) | 98.1% (96.1%–99.1%) | 96.0% (93.6%–97.5%) |
| Sep 2024 | 463 | 39 (8.4%) | 424 (91.6%) | 53.8% (38.6%–68.4%) | 98.1% (96.3%–99.0%) | 94.4% (91.9%–96.1%) |
| Oct 2024 | 478 | 58 (12.1%) | 420 (87.9%) | 70.7% (58.0%–80.8%) | 99.3% (97.9%–99.8%) | 95.8% (93.6%–97.3%) |
| Nov 2024 | 490 | 49 (10.0%) | 441 (90.0%) | 71.4% (57.6%–82.2%) | 98.2% (96.5%–99.1%) | 95.5% (93.3%–97.0%) |
| Dec 2024 | 454 | 50 (11.0%) | 404 (89.0%) | 62.0% (48.2%–74.1%) | 97.3% (95.2%–98.5%) | 93.4% (90.7%–95.3%) |
| Jan 2025 | 483 | 61 (12.6%) | 422 (87.4%) | 60.7% (48.1%–71.9%) | 98.6% (96.9%–99.3%) | 93.8% (91.3%–95.6%) |
| Feb 2025 | 434 | 48 (11.1%) | 386 (88.9%) | 64.6% (50.4%–76.6%) | 99.7% (98.5%–100.0%) | 95.9% (93.5%–97.4%) |

| | | | | | | |
|---|---|---|---|---|---|---|
| Mar 2025 | 483 | 49 (10.1%) | 434 (89.9%) | 55.1% (41.3%–68.1%) | 99.5% (98.3%–99.9%) | 95.0% (92.7%–96.6%) |
| Apr 2025 | 458 | 36 (7.9%) | 422 (92.1%) | 58.3% (42.2%–72.9%) | 99.5% (98.3%–99.9%) | 96.3% (94.1%–97.7%) |
| May 2025 | 482 | 48 (10.0%) | 434 (90.0%) | 64.6% (50.4%–76.6%) | 99.1% (97.7%–99.6%) | 95.6% (93.4%–97.1%) |
| Jun 2025 | 472 | 40 (8.5%) | 432 (91.5%) | 62.5% (47.0%–75.8%) | 99.8% (98.7%–100.0%) | 96.6% (94.6%–97.9%) |
| Jul 2025 | 522 | 51 (9.8%) | 471 (90.2%) | 68.6% (55.0%–79.7%) | 99.4% (98.1%–99.8%) | 96.4% (94.4%–97.7%) |
| Aug 2025 | 466 | 41 (8.8%) | 425 (91.2%) | 51.2% (36.5%–65.7%) | 99.3% (97.9%–99.8%) | 95.1% (92.7%–96.7%) |
| Sep 2025 | 467 | 53 (11.3%) | 414 (88.7%) | 58.5% (45.1%–70.7%) | 99.8% (98.6%–100.0%) | 95.1% (92.7%–96.7%) |
| Oct 2025 | 543 | 51 (9.4%) | 492 (90.6%) | 60.8% (47.1%–73.0%) | 98.6% (97.1%–99.3%) | 95.0% (92.9%–96.6%) |
| Nov 2025 | 530 | 55 (10.4%) | 475 (89.6%) | 67.3% (54.1%–78.2%) | 99.4% (98.2%–99.8%) | 96.0% (94.0%–97.4%) |
| Dec 2025 | 514 | 45 (8.8%) | 469 (91.2%) | 62.2% (47.6%–74.9%) | 99.1% (97.8%–99.7%) | 95.9% (93.8%–97.3%) |
| Jan 2026 | 566 | 47 (8.3%) | 519 (91.7%) | 72.3% (58.2%–83.1%) | 99.4% (98.3%–99.8%) | 97.2% (95.5%–98.3%) |
| Feb 2026 | 472 | 49 (10.4%) | 423 (89.6%) | 59.2% (45.2%–71.8%) | 99.8% (98.7%–100.0%) | 95.6% (93.3%–97.1%) |
| Mar 2026 | 269 | 25 (9.3%) | 244 (90.7%) | 48.0% (30.0%–66.5%) | 99.2% (97.1%–99.8%) | 94.4% (91.0%–96.6%) |

#### Table S4b: Quarterly temporal stability and performance drift

| Period | Total N | PE positive, No. (%) | PE negative, No. (%) | Sensitivity (95% CI) | Specificity (95% CI) | Accuracy (95% CI) |
|---|---|---|---|---|---|---|
| 2023 Q3 | 980 | 83 (8.5%) | 897 (91.5%) | 66.3% (55.6%–75.5%) | 98.0% (96.9%–98.7%) | 95.3% (93.8%–96.5%) |
| 2023 Q4 | 1,244 | 95 (7.6%) | 1,149 (92.4%) | 71.6% (61.8%–79.7%) | 98.3% (97.3%–98.9%) | 96.3% (95.1%–97.2%) |
| 2024 Q1 | 1,292 | 133 (10.3%) | 1,159 (89.7%) | 67.7% (59.3%–75.0%) | 98.4% (97.5%–98.9%) | 95.2% (93.9%–96.2%) |
| 2024 Q2 | 1,358 | 123 (9.1%) | 1,235 (90.9%) | 66.7% (57.9%–74.4%) | 98.2% (97.3%–98.8%) | 95.4% (94.1%–96.4%) |
| 2024 Q3 | 1,279 | 112 (8.8%) | 1,167 (91.2%) | 59.8% (50.6%–68.4%) | 98.3% (97.4%–98.9%) | 94.9% (93.6%–96.0%) |
| 2024 Q4 | 1,422 | 157 (11.0%) | 1,265 (89.0%) | 68.2% (60.5%–74.9%) | 98.3% (97.4%–98.8%) | 94.9% (93.7%–96.0%) |
| 2025 Q1 | 1,400 | 158 (11.3%) | 1,242 (88.7%) | 60.1% (52.3%–67.4%) | 99.3% (98.6%–99.6%) | 94.9% (93.6%–95.9%) |
| 2025 Q2 | 1,412 | 124 (8.8%) | 1,288 (91.2%) | 62.1% (53.3%–70.2%) | 99.5% (99.0%–99.8%) | 96.2% (95.1%–97.1%) |
| 2025 Q3 | 1,455 | 145 (10.0%) | 1,310 (90.0%) | 60.0% (51.9%–67.6%) | 99.5% (98.9%–99.7%) | 95.5% (94.3%–96.5%) |
| 2025 Q4 | 1,587 | 151 (9.5%) | 1,436 (90.5%) | 63.6% (55.7%–70.8%) | 99.0% (98.4%–99.4%) | 95.7% (94.5%–96.6%) |
| 2026 Q1 | 1,307 | 121 (9.3%) | 1,186 (90.7%) | 62.0% (53.1%–70.1%) | 99.5% (98.9%–99.8%) | 96.0% (94.8%–97.0%) |

**LLM Prompts for Reference Standards**

system_role = "You are a thoracic radiologist expert in pulmonary embolism (PE)."

prompt_3_rules = f"""Tasks (strict)

Using only the original report text (ignore any addendum content), determine whether the original report describes pulmonary embolism.

Critical decision rules:

- If the original report contains explicit, definitive wording such as “positive for PE”, “demonstrates pulmonary embolism,” “evidence of pulmonary embolism”, “segmental pulmonary embolism”, “subsegmental pulmonary embolism”, “Visualized filling defect,” “visualized thrombus,” “saddle pulmonary embolism,” or “Occlusive thrombus”, “could represent PE”, “may represent”, “mixing artifact vs thrombus”, then set original_PE_diagnosis = “True”
- If the original report contains explicit resolution wording such as “negative for pulmonary embolism”, “interval resolution,” “resolved,” “no new filling defect,” “resolution of”, “resolved bilateral”, “can not be excluded”, “suboptimal opacification”, “limited study”, “poor resolution”, “poor contrast”, “likely artifact”, then set original_PE_diagnosis = “False”
- Under no circumstances should you output “Indeterminate” as the final original_PE_diagnosis.

- Prefer impression or conclusion wording over findings if both are present.

Output

Return a single-line output only, using the exact keys:  ”True|False”

""" Prompt for chronicity and location:

system_role = "You are a thoracic radiologist expert in pulmonary embolism (PE)."

### The Prompts

prompt_chronicity = """ Task (strict)

(A). Using the report text provided, classify the age of the pulmonary embolus (PE) as acute, chronic, or age-indeterminate.

Critical decision rules (apply in this order)

1. Classify the PE as acute if the report contains the words "acute", "new", but do not consider negated statements such as "resolved acute PE", "no acute PE seen", "interval resolution of prior PE".
2. Classify the PE as chronic if the report contains “chronic,” "unchanged," “organized,” "persistent," “residual,” “recanalized,” “old,” “partial web,” “again noted,” ”stable”.
3. Classify the PE as age-indeterminate only when the report contains “age indeterminate” "uncertain,".
4. If a PE is described and no chronic or age-indeterminate descriptors are present, classify the PE as acute.
5. If more than one clot is described with different ages, apply global priority: acute > chronic > age-indeterminate

(B). Important

- Use the wording in the report exactly as written. Prefer the impression over the findings section, and use findings only if the impression does not provide sufficient information.
- Use only positive assertions of thrombus or embolus. Ignore negated statements.

Output

Return a single-line label only: Acute|Chronic|Age_Indeterminate.

"""prompt_location = """ Using the report text provided, classify the location of (most proximal) pulmonary arterial branch involved in the PE.

Hierarchy of priority: Central > Lobar_or_Segmental > Subsegmental

1. Classify as Central if the report explicitly describes thrombus in: the main pulmonary artery, the right pulmonary artery, the left pulmonary artery, the distal right or left pulmonary artery, the interlobar pulmonary artery, the truncus anterior, or describes a saddle pulmonary embolus.
2. Classify as Lobar_or_Segmental when the report explicitly describes thrombus in: a lobar pulmonary artery, a segmental pulmonary artery, a segmental branch, or any combination of lobar and segmental arteries. This includes cases where lobar and segmental emboli are both described. Do not infer this category from lobe-based wording alone (for example, “right lower lobe pulmonary embolism”) without vessel-level terminology.
3. Classify as Subsegmental when the report explicitly describes:” subsegmental pulmonary artery”, “subsegmental branches”.
4. Classify as indeterminate if the report mentions pulmonary embolism without specifying an arterial level, the report localizes PE only by lung lobe (for example, “right upper lobe pulmonary emboli”) without vessel-level terminology, vague distal or peripheral wording without specifying segmental or subsegmental arteries or insufficient information of the arterial level involved.

Important:

- Use the wording in the report exactly as written. Prefer impression over findings section and use findings only if impression does not provide sufficient information.
- Use only positive assertions of thrombus or embolus. Ignore negated statements.
- When multiple emboli are described at different arterial levels, select the most proximal level according to the hierarchy.

Output: Return a single-line labels only: Central|Lobar_Or_Segmental|Subsegmental|Indeterminate."""
